# Which came first, obstructive sleep apnea or hypertension? A retrospective study of electronic records over 10 years, with separation by sex

**DOI:** 10.1101/2020.01.27.20019018

**Authors:** Eunjoo An, Michael R. Irwin, Lynn V. Doering, Mary-Lynn Brecht, Karol E. Watson, Ravi S. Aysola, Andrea P. Aguila, Ronald M. Harper, Paul M. Macey

## Abstract

**Objectives:** Obstructive sleep apnea (OSA) is a risk factor for hypertension (HTN), but the clinical progression of the sleep disorder to the high blood pressure condition is unclear. There are also sex differences in prevalence, screening and symptoms of OSA. The objective was to estimate the time from OSA to HTN diagnoses, with sex-specific quantification.

**Design:** Retrospective analysis of electronic health records (EHR) over a 10-year period (2006 to 2015 inclusive).

**Setting:** UCLA Health System in Los Angeles, California, USA.

**Participants:** 4848 patients: female N=2086, mean [age±std] = 52.8±13.2 years; male N=2762, age=53.8±13.5 years. These patients were selected from 1.6 million patients with diagnoses in the EHR who met the criteria of: diagnoses of OSA and HTN; in long-term care defined by ambulatory visits at least one year prior and one year subsequent to the first OSA diagnosis; no diagnosis of OSA or HTN at intake; and a sleep study performed at UCLA.

**Primary and secondary outcome measures:** The primary outcome measure in each patient was time from the first diagnosis of OSA to the first diagnosis of HTN (in days). Since HTN and OSA are progressive disorders, a secondary measure was relationship between OSA-to-HTN time and age.

**Results:** The mean, std and 95% confidence intervals of the time from OSA to HTN diagnoses were: all - 732 ± 1094.9 [-764.6, -701.8] days; female -815.9 ± 1127.3 [-867.3, -764.2] days; and male -668.6 ± 1065.6 [-708.1, -626.8] days. Age was negatively related to time from OSA to HTN diagnosis in both sexes.

**Conclusions:** HTN was on average diagnosed years prior to OSA, with a longer separation in females. Our findings suggest under-screening of OSA, more so in females than males. Undiagnosed OSA may delay treatment for the sleep disorder and perhaps affect the development and progression of HTN.

**Brief Summary:**

- OSA is a risk factor for HTN so, in people with both conditions, the sleep disorder should typically precede the high blood pressure. However, the clinical sequence of these two conditions is unclear.
- For patients in long-term care in the UCLA health system, the diagnosis of OSA usually precedes by years the diagnosis of HTN in patients with both conditions. The later diagnosis of OSA versus HTN may reflect a lack of OSA screening for years after sleep disorder onset.

**Strengths and limitations of this study:**

- Approximately 5000 is a large enough sample to provide reliable effect size and confidence interval calculations.
- Inclusion criteria ensure observations are from patients likely to be in regular contact with the UCLA health system, and hence to be in a position to be regularly screened for HTN and OSA.
- Separation by sex highlights the presence of clinical differences in patients with HTN and OSA, although the nature of the differences (for example under-screening, or different progression in females and males) cannot be inferred.
- Limited generalizability due to data in a single health system and to the specific 10-year time period, the latter encompassing a change in diagnostic criteria for OSA.
- No data on diagnoses in other clinical settings or prior to 2006, meaning 1) many patients with HTN and OSA were likely excluded because the diagnoses were not recorded at UCLA and 2) of the included patients some may have had prior diagnoses thus affecting interpretation of the time from OSA to HTN measure.

## Introduction

Obstructive sleep apnea (OSA), a breathing disorder that effects around 30 million people nationally, is a primary risk factor for hypertension (HTN) ^2^. When comorbid with OSA, HTN is often treatment-resistant ^3^. An accepted model is that OSA pathophysiology leads to HTN ^4^, implying that treating OSA would help prevent or resolve HTN ^5^. However, randomized trials of treating OSA with continuous positive airway pressure (CPAP) show modest effects on HTN ^5^, confirming links between the two conditions are not simplistic. Factors such as the association between rapid-eye movement (REM) OSA and typical time time-of-night CPAP usage may explain the CPAP findings ^6^, but further data are needed to understand OSA-HTN relationships. Other relevant information is whether on average a diagnosis of OSA precedes a diagnosis of HTN. Despite the accepted pathophysiologic model, there is little clinical evidence on the question of whether OSA or HTN appear first in the clinical record. Even though there are reasons other than disease progression that could influence the timing of diagnoses (differences in HTN and OSA screening, multi-factorial nature of HTN), establishing the health record occurrence of HTN relative to OSA would contribute to our understanding of clinical and maybe mechanistic relationships between the conditions.

Assuming OSA is a precursor to HTN, we initially hypothesized that in patients with both conditions a diagnosis of OSA precedes a diagnosis of HTN. This hypothesis also assumes equivalent screening for both HTN and OSA, which, while unlikely to be true, nevertheless provides a starting point. Both HTN and OSA are progressive so clinical diagnoses usually occur months to years after condition onsets. Since screening for OSA is presumably less frequent than for HTN, the delay between onset and diagnosis likely differs between the two conditions, with OSA most likely having a longer delay than HTN ^5^. Hence, a finding that OSA precedes HTN clinically would strongly support the model of OSA preceding HTN; alternatively, if HTN precede OSA diagnoses, we could not rule out a longer delay to OSA screening. In either case, quantifying the time between diagnoses would be useful.

Our objective was to test the hypothesis that OSA precedes HTN using electronic health records (EHR) within the University of California, Los Angeles (UCLA) Health System. We assessed the time between OSA and HTN diagnoses in patients with both conditions. We also aimed to describe timing of diagnoses with respect to age and gender, since both factors influence prevalence and severity of both conditions ^7 8^.

## Methods

We evaluated EHR over a 10-year period beginning January 2006; the procedures were approved by the UCLA Institutional Review Board. The patient records containing diagnoses of HTN or OSA were retrieved. OSA was based on the following diagnostic codes: ICD-9 327.2, 327.20, 327.21, 327.2, 327.24 and 780.57; ICD-10 G47.30 and G47.33. HTN was based on: ICD-9 401.0, 401.1 and 401.9; ICD-10 I10. During the timeframe, the American Academy of Sleep Medicine scoring rules for respiratory events were updated ^9 10^; the UCLA Sleep Disorders Center transitioned from the 1999 to 2012 rules in the year following their release. Furthermore, UCLA transitioned from ICD-9 to ICD-10 In patients with both conditions, the time from first diagnosis of OSA to first diagnosis of HTN was calculated. The month and day of the first encounter date was converted to January 1 to remove personal identifiers (date of visit), but otherwise times between diagnoses were accurate to within a day. The day of first OSA diagnosis was set as 0 (relative starting point), and the time difference represented as days to first HTN diagnosis. Sleep study parameters were not available in the EHR.

We calculated descriptive statistics of time from OSA to HTN, including the mean which we tested for being non-zero with a 1 sample t-test. The relationship between age and time from OSA to HTN was calculated using a linear regression. We repeated these analyses for females and males separately. The location of diagnoses (outpatient vs inpatient) within the 10-year period affects possible OSA to HTN times, so we viewed these times separately by year and by gender. Statistics were calculated with MATLAB.

## Results

Of the 1.6 million patient records evaluated (n = 1,654,067) with at least one diagnosis, approximately 2% (29,764) contained OSA diagnoses and 14% (192,771) contained HTN diagnoses. There were 16,974 (1%) patient records with both diagnoses of OSA and HTN, of which 36% (6124) had a sleep study within the UCLA Health System (A sleep study is required to diagnose OSA). We defined long-term care as patients with encounters at least a year before and after OSA diagnosis, who comprise 29% (4848) of records the OSA/HTN patients. Data are available online ^1^.

Table 1 shows characteristics of OSA and HTN diagnoses and patient demographics, separated by gender. Hypertension was typically diagnosed earlier than OSA (*P* < 0.05; mean time difference = -732 days; median = -532 days). Females showed earlier HTN diagnosis than males (*P* < 0.05 female vs. male; days delay: females mean = -815.9 days/median = -610, males mean = -668.6 days/median = -451). Age was negatively related to time from OSA to HTN diagnosis in both sexes (*P* < 0.05). These findings are illustrated in Figures 1 and 2.

**Table 1.**
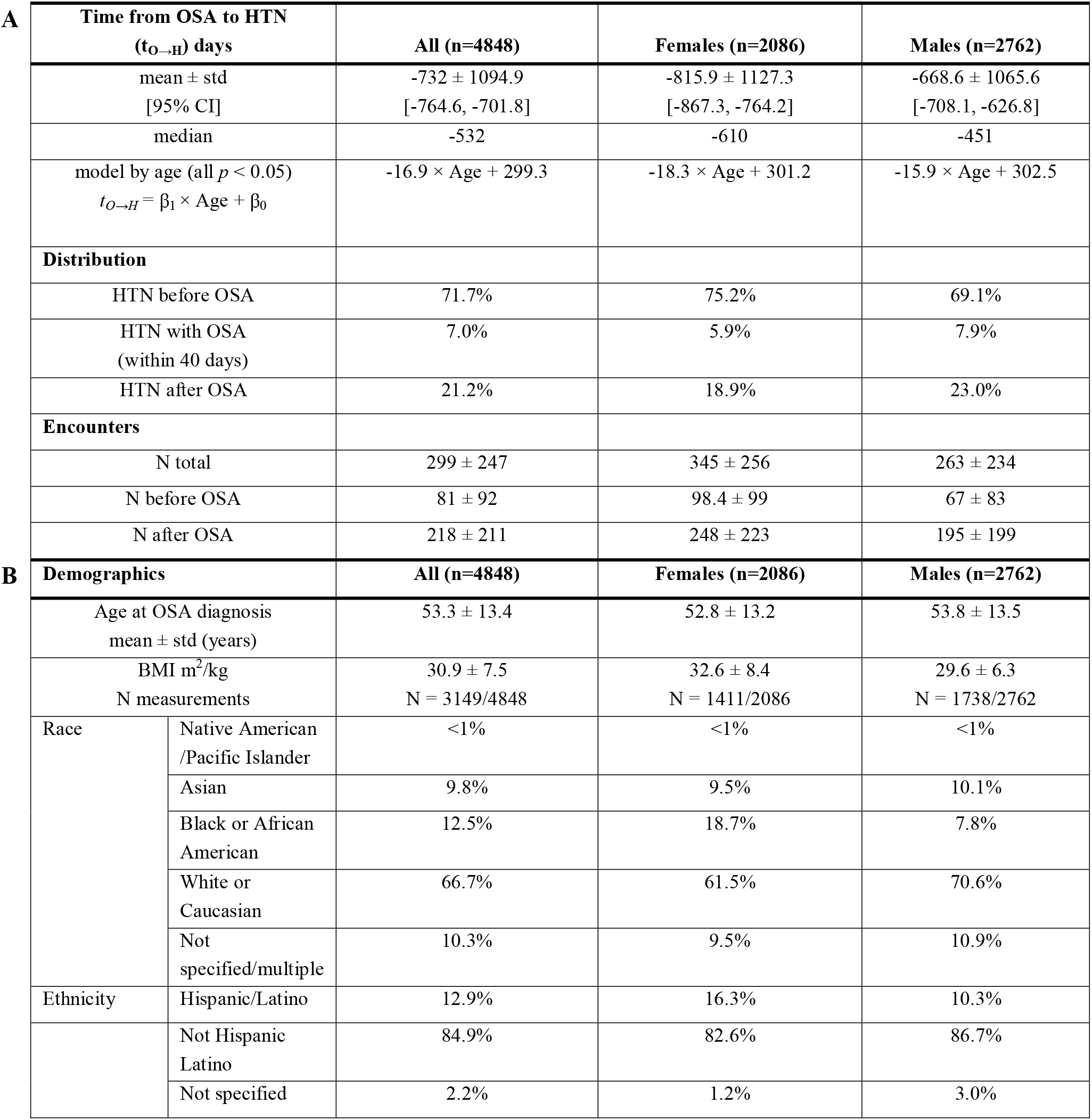
Patient characteristics separated by gender. **A**. Days from first OSA to first HTN diagnosis (*t*_*O*→*H*_;) mean and 95% confidence intervals [CI], median, and regression model by age. **B**. Demographics. Approximate age at time of OSA diagnosis. BMI was the closest recorded value to the date of OSA diagnosis; N indicates subset of patients with BMI values. Race and ethnicity were as recorded in EHR.

**Figure 1.**
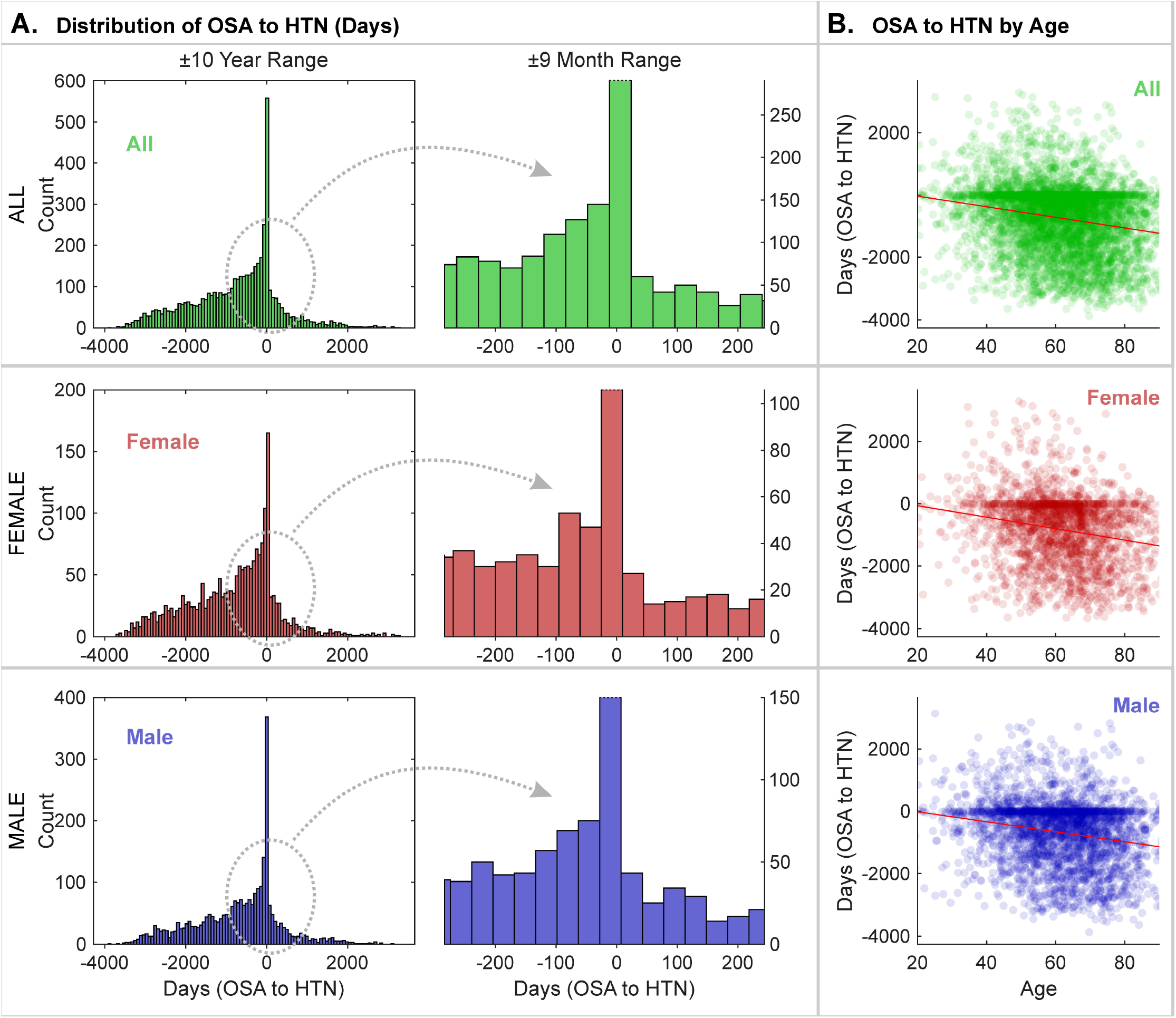
Time from OSA to HTN: Distribution and time by age, separated by sex. **A**. Distribution of time from OSA to HTN diagnoses, for combined (“ALL”) and female and male patients; ±10 year and ±9 month ranges are shown. **B**. Scatterplots of time from OSA to HTN diagnoses with respect to age at OSA diagnosis, for combined (“ALL”) and female and male patients.

**Figure 2.**
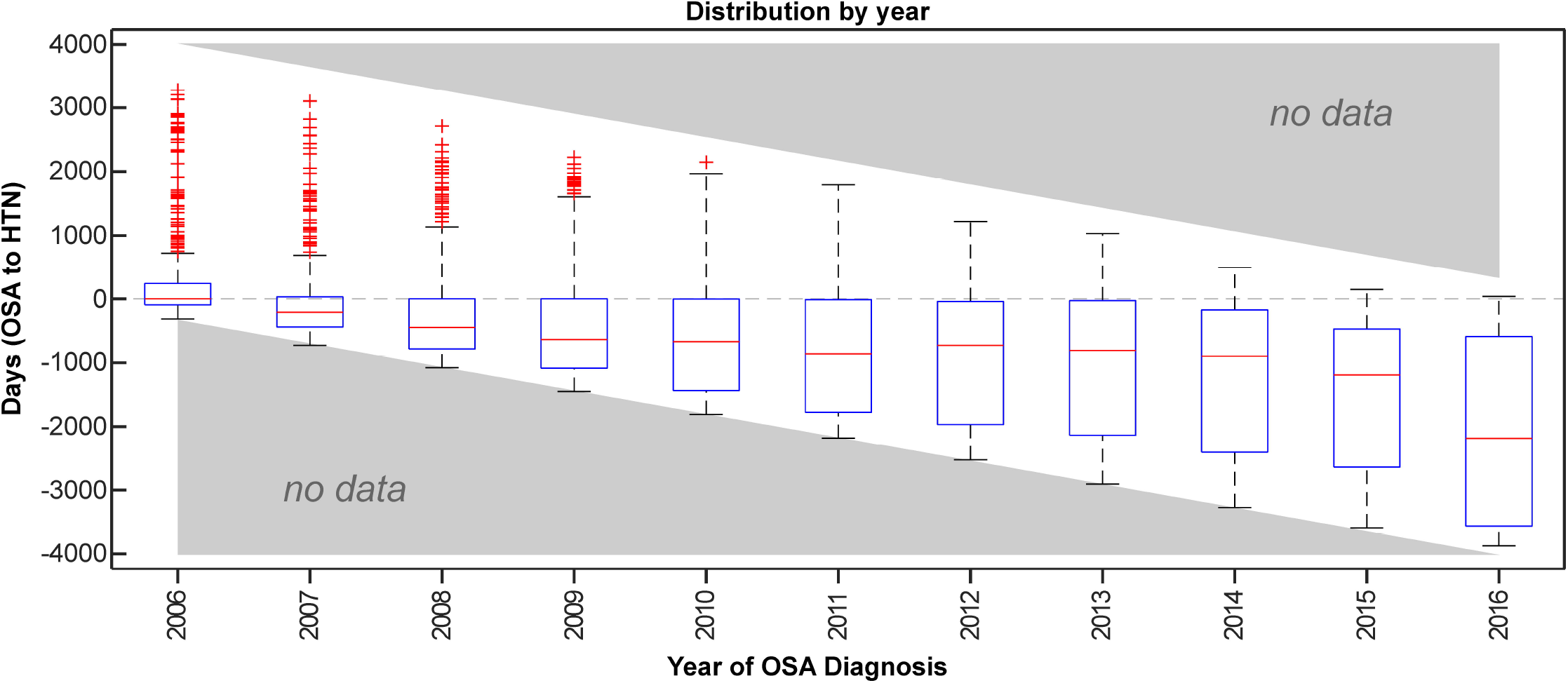
Distribution by year of OSA diagnosis in patients with OSA and HTN. Distribution by year of time from OSA to HTN diagnoses, with box plots by year of OSA diagnosis (box is between 25^th^ and 75^th^ percentiles, red line is median, whisker lines are ranges with outliers defined as more than one interquartile range from box marked as red “+”). Grey areas indicate no data are available for diagnoses prior to 2006 or after 2015 (so a delay from before 2006 or after 2015 could not be observed).

We repeated these analyses for the 16,974 records of all patients with HTN and OSA diagnoses and the 6124 with a sleep study, and the direction of effects did not change for either set.

## Discussion

From 2006 to 2016 in the UCLA Health System, HTN was on average diagnosed years prior to OSA, with a longer separation in females. The time between diagnoses could reflect differences in screening, specifically that HTN is more frequently screened than OSA. Assuming the OSA-causes-HTN model is correct, these findings would reflect a delay in OSA relative to HTN screening of several years. An alternative possibility is that OSA is not a principal mechanism of HTN–the findings could reflect both conditions co-occurring, or that HTN may precede the development of OSA due to other factors. The present data cannot distinguish between these possibilities. Practically speaking, a combination of effects is likely at play, since there is more than one cause of HTN, just as OSA is a heterogenous disorder.

Considering possible common factors, the co-occurrence of OSA and HTN may be associated with structural and biological effects of obesity, which is highly prevalent in both conditions ^4 11^. In OSA, the extra layer of adipose tissue surrounding the neck may contribute to the narrowing of the upper airway, and rigidity of the thoracic and abdomen walls due to the adipose tissue deposits surrounding tissues, further compromising the air flow ^12^. In HTN, an increase in adipose tissue, especially surrounding the abdomen wall, may contribute to progression of HTN due to excess amount of adipokines produced ^13 14^. Adipokines play a key role in body homeostasis, including insulin regulation, lipid and glucose metabolism, coagulation, and angiogenesis and vascular remodeling, which all affect blood pressure regulation ^14^.

The longer separation seen in females may be partially explained by more frequent screening in males due to differences in clinical presentation between the genders. For instance, women typically report fatigue, insomnia, and depression, symptoms not classically associated with OSA, whereas, men typically report daytime sleepiness, the defining symptom of the sleep disorder ^15^. Sex differences in OSA, such as structural changes in the brain^16^ and menopause,^17^ mean that the OSA-HTN relationship may differ between males and females. Pre-menopausal women and postmenopausal women on hormone replacement therapy have shown significantly lower OSA prevalence, suggesting hormones may have a protective role women ^18^. Other major differences between the sexes are that men have a higher prevalence of OSA than women by a factor of 2:1 ^11^, which could influence the likelihood of screening based on expectations that males are more likely to have the sleep condition.

While these findings are from one health system and may not generalize, our analysis could be replicated with other EHR datasets. The analysis could also be replicated in future time periods, since screening for OSA may improve with time. OSA is a relatively recently discovered disease, and awareness has been an increasing over the past two decades ^11^. Similarly, screening for HTN, perhaps with home testing, may also lead to earlier diagnosis relative to onset, so the patterns seen from 2006 to 2016 may well differ in the future. Additionally, the present data and the refractory nature of HTN in CPAP-treated OSA leave open the possibility that the model of OSA as principal mechanism of high blood pressure is complex and may warrant consideration of additional factors such as obesity and other comorbid conditions, or even bi-directional influences ^19^.

## Data Availability

Data are publicly available in an online repository.

https://dataverse.harvard.edu/dataset.xhtml?persistentId=doi:10.7910/DVN/ZY1LDT

